# Time-to-Statin Prescription for Primary Atherosclerotic Cardiovascular Disease Prevention in a Lung Cancer Screening Program in Missouri: A Retrospective Cohort Study

**DOI:** 10.64898/2026.09.16.26363270

**Authors:** Isaac Che Ngang, Eyerusalem Zewde, Sridharan Gopalsamy Ramaswamy, Akila Anandarajah, Benjamin Bowe, Beryne Odeny

## Abstract

**Background:** Individuals undergoing lung cancer screening (LCS) represent a population at high risk for atherosclerotic cardiovascular disease (ASCVD), yet opportunities for cardiovascular prevention during screening encounters may be underutilized. While prior studies have examined whether statins are prescribed in eligible patients, little is known about the *timing* of statin initiation following LCS, particularly among statin-naïve individuals.

**Methods:** We conducted a retrospective cohort study using electronic health record data from a large academic health system in Missouri. Adults aged 50–80 who underwent LCS between January 1, 2015, and December 31, 2023, were statin-naïve, and met 2019 ACC/AHA criteria for primary prevention were included. The primary outcome was time-to-statin initiation following LCS. Kaplan–Meier methods and Cox proportional hazards models were used to evaluate timing and predictors of statin initiation across demographic, clinical, and socioeconomic subgroups.

**Results:** Among 3,100 statin-eligible, statin-naïve individuals who had undergone LCS, only 27.3% were prescribed a statin within one year of LCS. Uptake accrued gradually (10.5% by 90 days; 17.8% by 180 days; 23.2% by 270 days; 27.1% by 360 days). In adjusted models, earlier statin initiation was independently associated with a higher ASCVD risk category, a cardiology visit in the year preceding LCS, and former (versus current) smoking; older age and male sex were associated with slower initiation. Race, insurance type, and area deprivation were not independently associated with time-to-statin initiation.

**Conclusions:** Despite high ASCVD risk, most statin-eligible patients undergoing LCS did not receive timely statin therapy. Earlier initiation tracked calculated ASCVD risk and specialty (cardiology) contact rather than race, sex, insurance, or area deprivation. Because most patients at high calculated risk still went untreated, integrating cardiovascular risk assessment and preventive decision support into LCS workflows may help reduce missed opportunities for ASCVD prevention.

## Background

Lung cancer remains a leading cause of cancer-related mortality in the U.S., with an estimated 650 new diagnoses and nearly 350 deaths occurring daily.^1^ Simultaneously, cardiovascular disease (CVD) is the leading cause of mortality in the U.S., accounting for ∼2,300 deaths each day.^2^ These conditions share smoking as a risk factor and those eligible for lung cancer screening (LCS) often present with hypertension and diabetes, and CVD remains the leading cause of mortality among those affected.^3^ Moreover, LCS frequently detects coronary artery calcification (CAC), a strong predictor of future ASCVD events that has been incorporated into standardized reporting frameworks as a prompt for ASCVD risk assessment and intervention.^4^ Given the clinical overlap between lung cancer and ASCVD, LCS provides an opportunistic setting for physicians to assess ASCVD risk and address gaps in preventive care.^5^ What remains unknown is how promptly statin-eligible, previously untreated patients initiate therapy following LCS, and whether that timing differs across clinical and sociodemographic subgroups the gap this study addresses.

Historically, the integration of ASCVD prevention into LCS workflows has been limited. The 2019 American College of Cardiology (ACC)/American Heart Association (AHA) guidelines recommend statin therapy for high-risk individuals, including smokers, yet this is often under-implemented in practice.^5^ Many LCS-eligible individuals come from underserved populations facing structural barriers to healthcare, such as lack of insurance, transportation, and consistent primary care access. These challenges disproportionately affect racial and ethnic minorities, particularly Black patients, who are less likely to receive statins despite meeting guideline criteria.^6^ If LCS, which requires multiple visits, specialist coordination, and complex imaging workflows can be successfully implemented in these populations, ASCVD risk assessment, a comparatively simpler and more streamlined process, should be even more accessible within the same encounter.

While many studies have focused on whether eligible individuals receive a statin prescription, far less have asked when statins are prescribed, a critical factor reflecting timely responsive interventions.^7,8^ Fewer studies have examined whether timing varies across demographic lines such as sex, race, place (rurality) and area deprivation, despite well-documented disparities in both cancer and ASCVD care.^5,9^

This study aims to assess whether the LCS process serves as a catalyst or a missed opportunity for ASCVD prevention in populations that have accessed LCS without prior primary ASCVD preventive care. By analyzing time-to-statin initiation following LCS, we seek to uncover whether a temporal relationship exists between LCS and lifesaving statin-prescription. A shorter time to initiation could suggest that LCS functions as an effective entry point for ASCVD risk management, whereas delays or absence of prescription may highlight enduring care gaps and structural barriers within the healthcare system. Our primary objective is to determine the association between LCS and time-to-statin prescription, correlates of time-to-statin prescription, and associated disparities (Figure 1).

**Figure 1.**
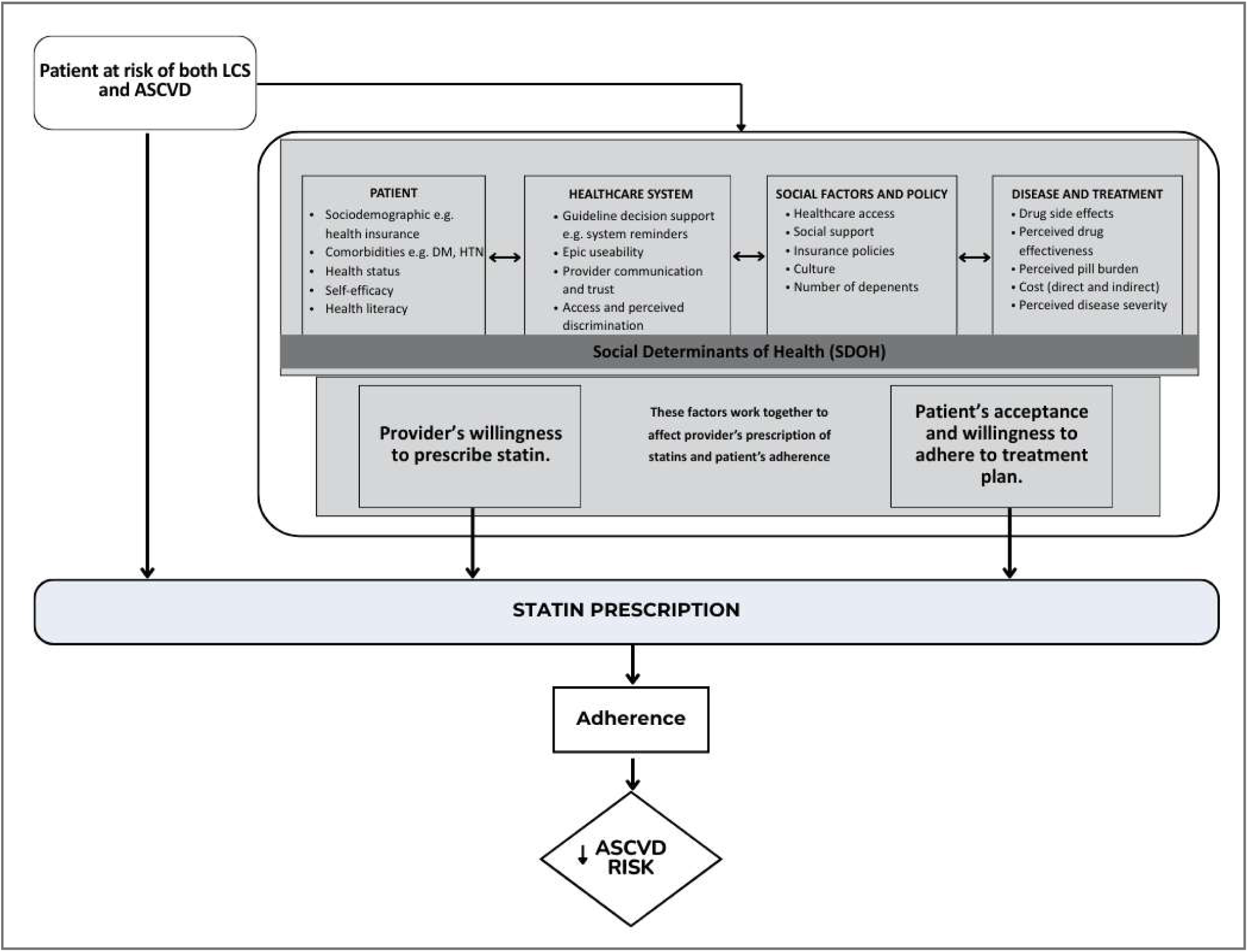
Conceptual framework

## Methods

### Study Design and Setting

This retrospective cohort study was conducted at Washington University in St. Louis School of Medicine, using electronic health record (EHR) data from the Barnes-Jewish Healthcare system and the Siteman Cancer Center. The study population included individuals who underwent LCS between January 1, 2015, and December 31, 2023. This study was designed and reported in accordance with the Strengthening the Reporting of Observational Studies in Epidemiology (STROBE) guidelines for observational research^10^; the completed STROBE checklist is provided as a supplementary file.

### Study Population

Eligible participants were adults aged 50 to 80 with a documented smoking history of at least 20 pack-years and no prior statin or statin-equivalent use at the time of their LCS. Patients were required to have available EHR data detailing ASCVD risk factors and medication history. Individuals were excluded if they were previously prescribed statins, lacked key clinical data (e.g., lipid profiles, smoking history, or prescription records), or did not complete an LCS during the study period (see Figure 3 for the study flowchart).

### Data Collection

Clinical data were extracted from Epic, the EHR system used by Barnes-Jewish Healthcare. Collected variables included patient demographics (age, sex, race/ethnicity, and socioeconomic status indicators), smoking history (pack-years and current or former smoking status), and relevant clinical parameters (blood pressure, lipid profiles, body mass index [BMI], and comorbidities such as hypertension and diabetes mellitus). ASCVD risk scores were calculated using the Pooled Cohort Equations as described by Goff et al.^11^ Where available, pre-calculated ASCVD risk scores documented in the EHR were used. For patients without a pre-existing score, risk was calculated using the most recently available clinical data. Correlation between pre-calculated and study-derived scores was assessed as part of quality control; accordingly, ASCVD scores in this dataset may not perfectly reflect values at the exact time of the first LCS encounter.

For statin therapy, we included all patients who were prescribed statins or statin-equivalent lipid-lowering medications, including ezetimibe, fibrates, and bile acid sequestrants. Throughout the remainder of the manuscript, these medications will be collectively referred to as “statins” for simplicity and consistency in terminology. Statin prescription data included the date of initiation, statin type and dosage, and whether the prescription occurred before or after LCS. Socioeconomic status was assessed using the Area Deprivation Index (ADI), a validated composite index developed by the University of Wisconsin–Madison, which measures neighborhood-level disadvantage using 17 indicators from the U.S. Census and American Community Survey, encompassing domains such as income, education, employment, and housing quality.^9^ Rurality was characterized using Rural–Urban Commuting Area (RUCA) codes, which classify census tracts according to population density, urbanization, and daily commuting patterns. RUCA codes were collapsed into four categories spanning metropolitan (category 1) to rural (category 4).

### Statin Eligibility Based on 2019 ACC/AHA Guidelines

Statin eligibility was determined according to the 2019 ACC/AHA guidelines for the primary prevention of CVD (Figure 2). Patients were considered eligible for statin therapy if they met any of the following criteria: (1) age 40–75 with diabetes and LDL-C between 70 and 189 mg/dL; (2) age 40–75 without diabetes, with LDL-C between 70 and 189 mg/dL, and an estimated 10-year ASCVD risk of 7.5% or higher based on the Pooled Cohort Equations; or (3) presence of risk-enhancing factors in patients with a borderline (5% to <7.5%) or intermediate (7.5% to <20%) 10-year ASCVD risk (Arnett et al., 2019). Risk-enhancing factors assessed included: family history of premature ASCVD, persistently elevated LDL-C ≥160 mg/dL, chronic kidney disease (CKD), chronic inflammatory conditions (e.g., rheumatoid arthritis, Systemic lupus erythematosus(SLE), HIV), and current smoking status.

**Figure 2.**
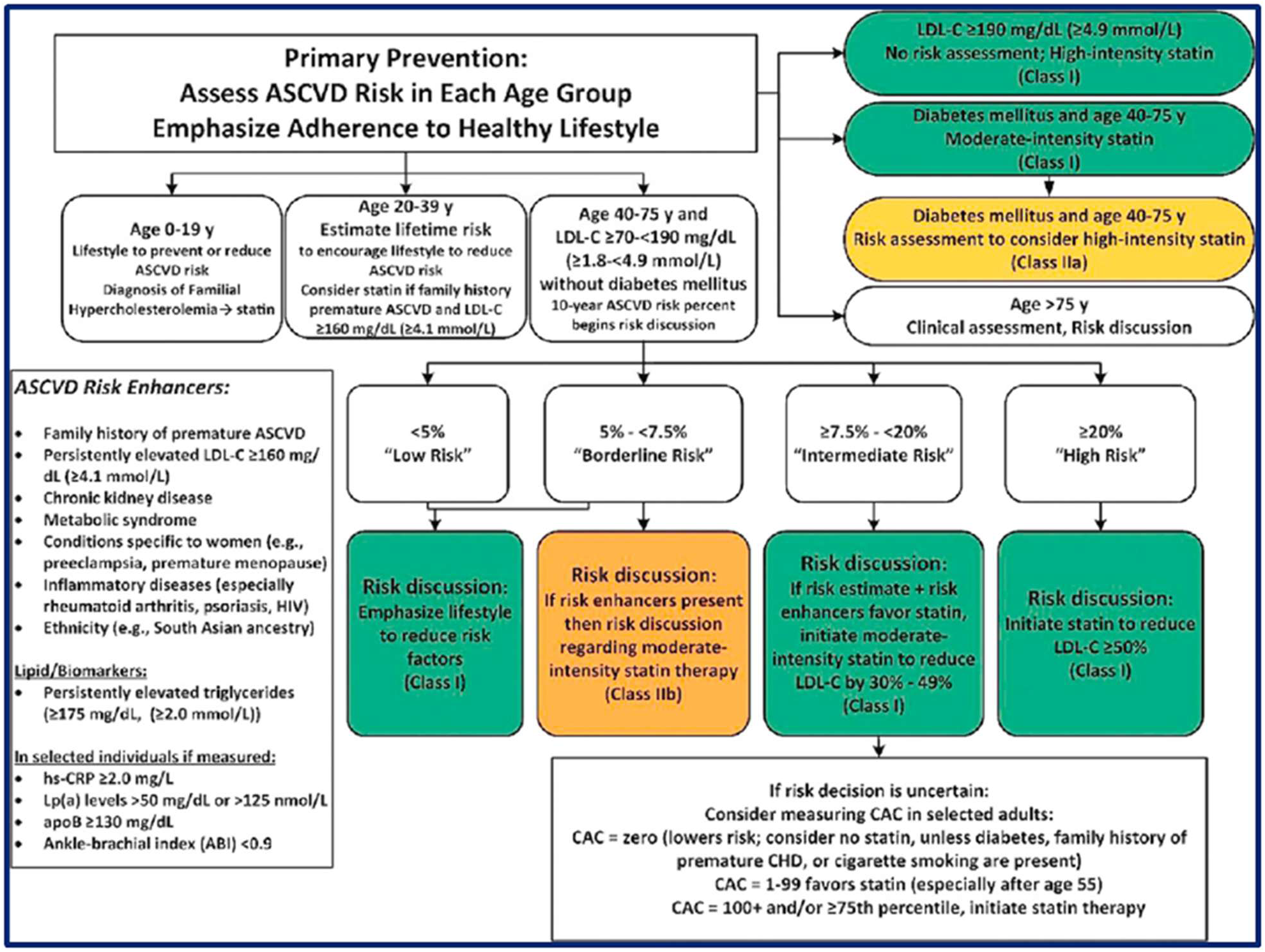
Statin eligibility following the 2019 ACC/AHA guidelines (Arnett et al., 2019).

**Figure 3:**
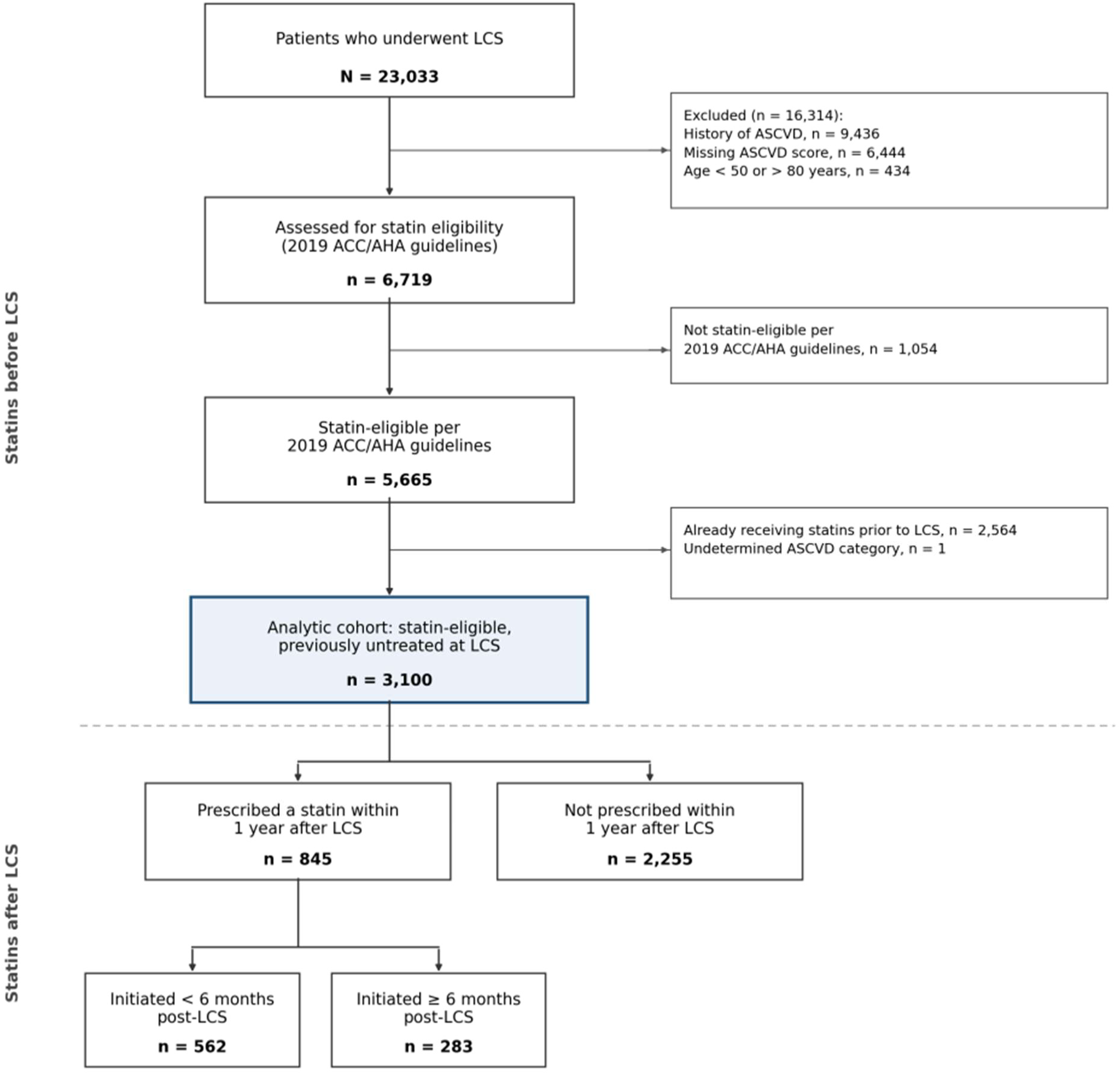
Study flowchart

### Outcomes

The primary outcome was the time-to-statin initiation relative to the first documented LCS, examined across race, sex, ASCVD risk categories, and levels of socioeconomic deprivation as measured by ADI. The secondary outcome was the identification of predictors associated with statin prescription within one year following LCS among patients who were statin-naïve at the time of screening.

### Data Analysis

Descriptive statistics were used to summarize patient characteristics, including sociodemographic variables, clinical risk factors, and prescription patterns. Chi-square tests were used to compare categorical variables across statin prescription status groups, including prescription rates by race, sex, insurance type, smoking status, comorbidity, and ASCVD risk category. T-tests or Mann-Whitney U tests were applied for continuous variables including age, BMI, and ASCVD risk scores across the same statin prescription status groups.

Kaplan-Meier survival analysis was conducted to assess time-to-statin initiation following LCS. Survival curves were stratified by ASCVD risk level, race, sex, and ADI quartile. For all Kaplan-Meier analyses, we assessed differences in time-to-statin initiation across subgroups using the log-rank test. In instances where survival curves crossed indicating a possible violation of the proportional hazards assumption the Breslow (Generalized Wilcoxon) test was additionally reported; however, crossing curves may reflect differential early event rates rather than a sustained difference, and Breslow results should be interpreted with this context in mind. Cox proportional hazards models were used to evaluate predictors of time-to-statin initiation following LCS, consistent with consistent with the time-to-event (survival analysis) framework used for the Kaplan–Meier analyses above. This approach supersedes the use of binomial logistic regression for this primary time-to-event outcome.

Missing data were present primarily in lipid profiles and ASCVD risk scores; patients missing an ASCVD score were excluded from the analytic cohort because ASCVD scores were required for eligibility determination. Among the final analytic cohort of 3,100 participants, missingness was modest for the remaining variables (e.g., packs of cigarettes per day: 15.6% missing; BMI: 4.5% missing). Multivariable models used complete-case analysis, and the sensitivity analysis described above confirmed the robustness of primary findings. A two-sided p-value <0.05 was considered statistically significant for all analyses. Data analysis was performed using R.

## Results

### Baseline Characteristics of the Study Cohort

A total of 23,033 patients underwent LCS between January 1, 2015, and December 31, 2023. After excluding patients with a history of ASCVD (n = 9,436), incomplete medical records (n=6,444 missing ASCVD scores), older than 80 or younger than 40(n=434), ineligible for statin therapy per 2019 ACC/AHA guidelines (n = 1,054), and already on statins at the time of LCS (n = 2,564), a total of 3100 met the inclusion criteria and were included in the final analytic cohort (figure 3).

The mean age of included participants was 66.2 years (SD 6.6), with 47.0% identifying as female. The racial composition included 73.7% White, 24.5% Black, and 1.8% identifying as Asian or other. Most patients were former smokers (59.3%), and common comorbidities included hypertension (71.5%), diabetes mellitus (29.9%), and chronic kidney disease (14.8%; Table 1).

**Table 1.** Baseline characteristics of the study population stratified by statin prescription status post-LCS.

| Characteristic | No statin<br>2,255 (72.7%) | Statin<br>845 (27.3%) | Total |
| --- | --- | --- | --- |
| **Age, years** | 65.9 [61.4–70.9]<br>mean 66.1 (6.6) | 66.6 [61.8–71.4]<br>mean 66.6 (6.6) | 3100 |
| **Sex** |  |  |  |
| Female | 1041 (71.4%) | 416 (28.6%) | 1457 |
| Male | 1214 (73.9%) | 429 (26.1%) | 1643 |
| **Race** |  |  |  |
| White | 1677 (73.4%) | 607 (26.6%) | 2284 |
| Black | 537 (70.6%) | 224 (29.4%) | 761 |
| Asian | 19 (67.9%) | 9 (32.1%) | 28 |
| Other | 22 (81.5%) | 5 (18.5%) | 27 |
| **Insurance type** |  |  |  |
| Medicare | 1011 (71.5%) | 402 (28.5%) | 1413 |
| Medicaid | 5 (83.3%) | 1 (16.7%) | 6 |
| Multiple | 581 (71.5%) | 232 (28.5%) | 813 |

| **Characteristic** | **No statin**<br>**2,255 (72.7%)** | **Statin**<br>**845 (27.3%)** | **Total** |
| --- | --- | --- | --- |
| None | 10 (90.9%) | 1 (9.1%) | 11 |
| Private | 648 (75.6%) | 209 (24.4%) | 857 |
| **RUCA category** |  |  |  |
| 1 | 1957 (72.9%) | 726 (27.1%) | 2683 |
| 2 | 168 (68.9%) | 76 (31.1%) | 244 |
| 3 | 94 (74.6%) | 32 (25.4%) | 126 |
| 4 | 36 (76.6%) | 11 (23.4%) | 47 |
| **ADI category** |  |  |  |
| 1 | 372 (69.9%) | 160 (30.1%) | 532 |
| 2 | 439 (73.3%) | 160 (26.7%) | 599 |
| 3 | 471 (74.5%) | 161 (25.5%) | 632 |
| 4 | 454 (71.6%) | 180 (28.4%) | 634 |
| 5 | 519 (73.8%) | 184 (26.2%) | 703 |
| **Smoking status** |  |  |  |
| Current | 956 (75.8%) | 306 (24.2%) | 1262 |
| Former | 1299 (70.7%) | 539 (29.3%) | 1838 |
| **Packs per day** | 0.3 [0.0–1.0]<br>mean 0.5 (0.7) | 0.0 [0.0–0.9]<br>mean 0.4 (0.5) | 2617 |
| **Cardiology visit** |  |  |  |
| No | 1813 (74.1%) | 635 (25.9%) | 2448 |
| Yes | 442 (67.8%) | 210 (32.2%) | 652 |
| **ASCVD risk category** |  |  |  |
| Borderline | 215 (79.9%) | 54 (20.1%) | 269 |
| Intermediate | 1325 (77.6%) | 382 (22.4%) | 1707 |
| High | 715 (63.6%) | 409 (36.4%) | 1124 |
| **ASCVD 10-yr score** | 14.9 [10.2–22.6]<br>mean 17.9 (10.8) | 19.6 [12.4–29.3]<br>mean 22.3 (13.0) | 3100 |

Values are n (%) within row unless otherwise noted. Continuous variables shown as median [IQR] and mean (SD).

### Baseline Characteristics of the Study Population by Statin Prescription Status

Among the 3,100 statin-eligible patients included in the analytic cohort, 27.3% (n = 845) had a statin prescription within one year following LCS (Table 1), while 72.7% (n = 2,255) did not. Patients with a statin prescription were slightly older than those without (median 66.6 vs. 65.9 years, p = 0.047). There were no statistically significant differences by sex (p = 0.14) or race (p = 0.29); the statin prescription rate was 29.4% among Black patients and 26.6% among White patients. Insurance type was not significantly associated with statin prescription (p = 0.12).

Former smokers were more likely to have a statin prescription than current smokers (29.3% vs. 24.2%, p = 0.002). Cardiology visits were strongly associated with statin prescription, with 32.2% of those with at least one cardiology visit in the preceding year having a statin prescription, compared with 25.9% among those without such a visit (p = 0.002). Patients with comorbid conditions including hypertension (30.4% vs. 19.3%, p < 0.001), diabetes mellitus (35.9% vs. 23.6%, p < 0.001), and CKD (35.3% vs. 25.9%, p < 0.001) were more likely to have a statin prescription. There was no significant association between HIV status and statin prescription (p = 0.71).

Rural–Urban Commuting Area (RUCA) categories, a measure of rurality, were not significantly associated with statin prescription status (p = 0.48), nor was area deprivation (ADI; p = 0.40).

Patients with one or more ASCVD risk-enhancing factors were more frequently prescribed statins than those without (28.6% vs. 23.8%, p = 0.007). BMI was higher in the statin group (30.1 vs. 28.8 kg/m², p < 0.001). As expected, ASCVD risk was strongly associated with statin prescription: prescription rates were 36.4% in the high-risk category, 22.4% in the intermediate-risk category, and 20.1% in the borderline-risk category; the overall difference across categories was statistically significant (p < 0.001). Additional details on baseline characteristics by statin prescription status are presented in Table 1.

Values are n (%) within row unless otherwise noted. Continuous variables shown as median [IQR] and mean (SD].

### Time-to-statin prescription post LCS

The median time-to-statin initiation was not reached within the follow-up period, as fewer than 50% of participants were prescribed a statin post-LCS. Due to the high level of censoring (72.7%), the mean time to prescription is not presented, as it may not accurately reflect the distribution of time to event. Cumulative statin initiation was 10.5% by day 90, 17.8% by day 180, 23.2% by day 270, and 27.1% by day 360, reaching 27.3% within one year.

### Time-to-statin prescription across clinical and demographic subgroups

Kaplan–Meier survival analysis was conducted to assess time-to-statin initiation following LCS across demographic and clinical subgroups. Due to high levels of censoring, the cumulative incidence at the end of follow-up is used as the primary descriptor, as median survival time was not reached in most subgroups and therefore has limited interpretive value.

There was a significant difference in time-to-statin initiation based on whether patients had a cardiology visit in the year preceding LCS (log-rank p < 0.001; Breslow p < 0.001). Patients with a prior cardiology visit experienced significantly shorter time-to-statin prescription compared to those without such a visit (Figure 4). Median time-to-statin initiation was not reached in either group due to censoring.

**Figure 4.**
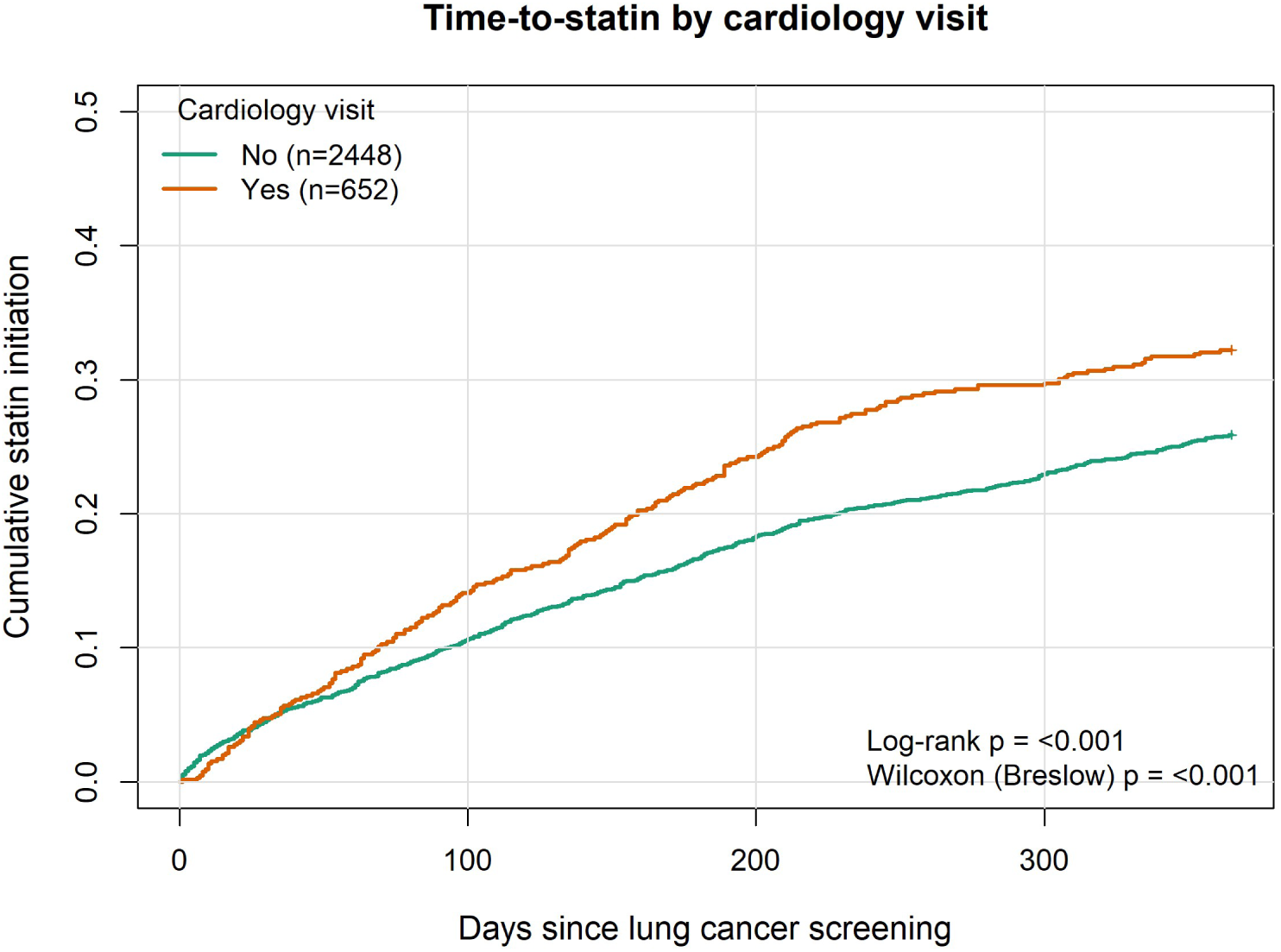
Time-to-statin prescription post-LCS by cardiology visit history in year preceding LCS

Former smokers began statins more quickly (log-rank *p* = 0.005) and had consistently shorter time-to-statin prescription compared with current smokers throughout the follow-up period (Figure 5). Despite these differences, the median time-to statin initiation was not reached in either group due to high censoring.

**Figure 5.**
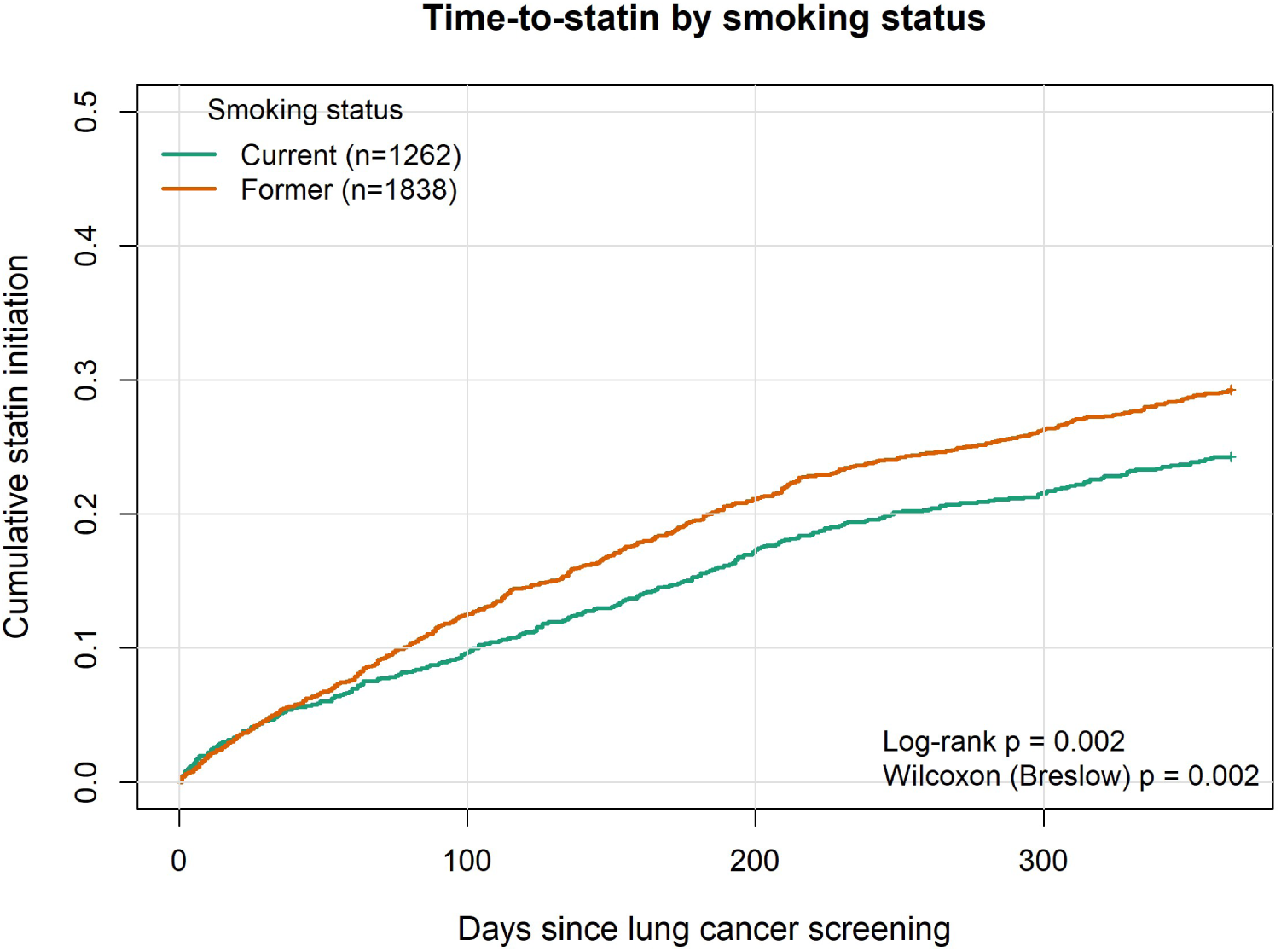
Time-to-statin prescription post-LCS by smoking status

ASCVD risk category was strongly associated with time-to-statin initiation (log-rank p < 0.001; Breslow p < 0.001). High-risk patients initiated statins substantially earlier and more frequently than intermediate- or borderline-risk patients, with clear and sustained separation of the curves (Figure 6); formal Cox model results are presented in the following section.

**Figure 6.**
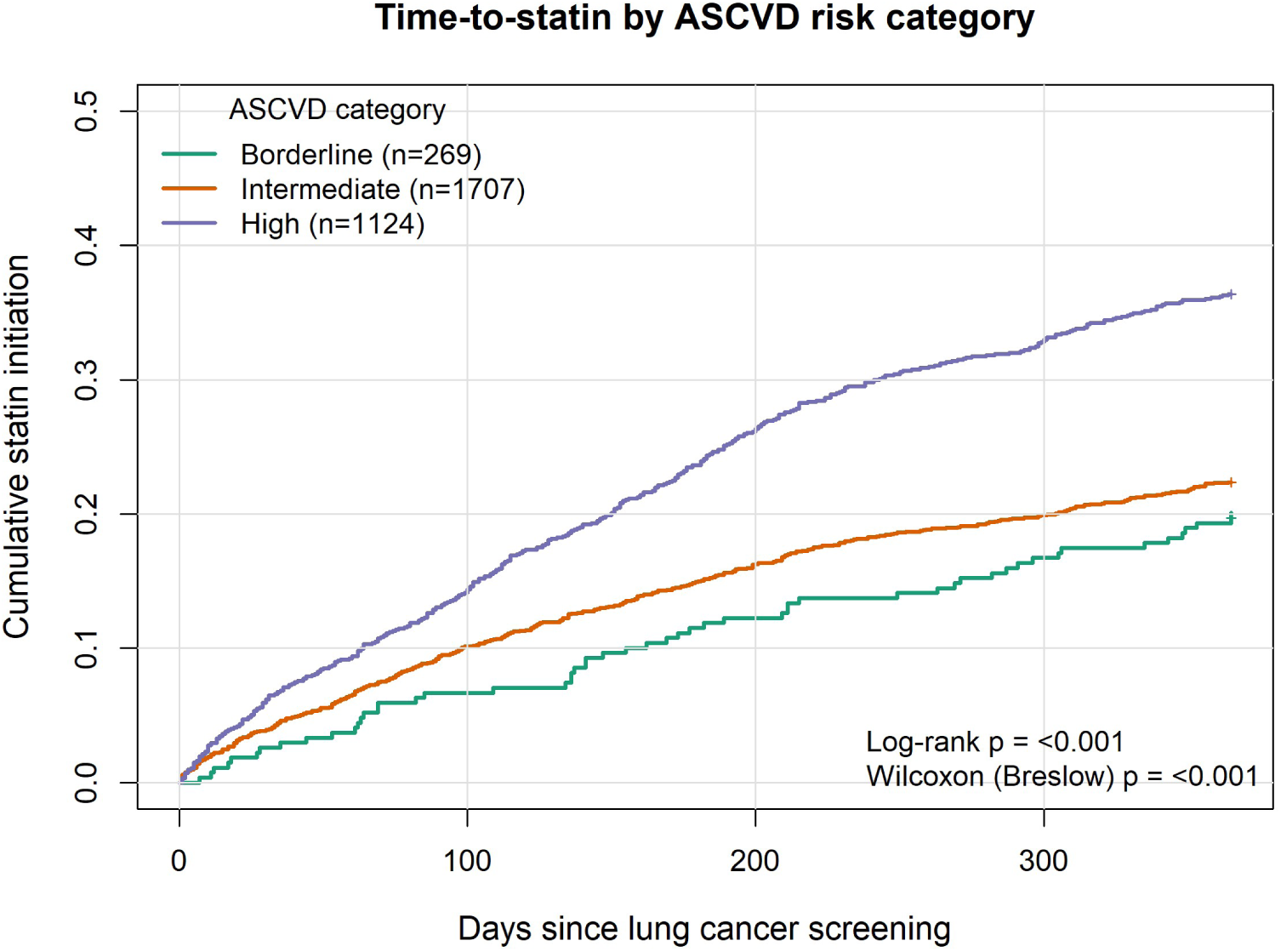
Time-to-statin prescription post-LCS by ASCVD risk categories

Statin initiation did not differ significantly by insurance type (log-rank p = 0.098; Breslow p = 0.075). Patients with Medicare, multiple insurance types, and private coverage had broadly similar trajectories, whereas the small Medicaid (n = 6) and no-insurance (n = 11) groups had sparse events and unstable estimates (Figure 7).

**Figure 7.**
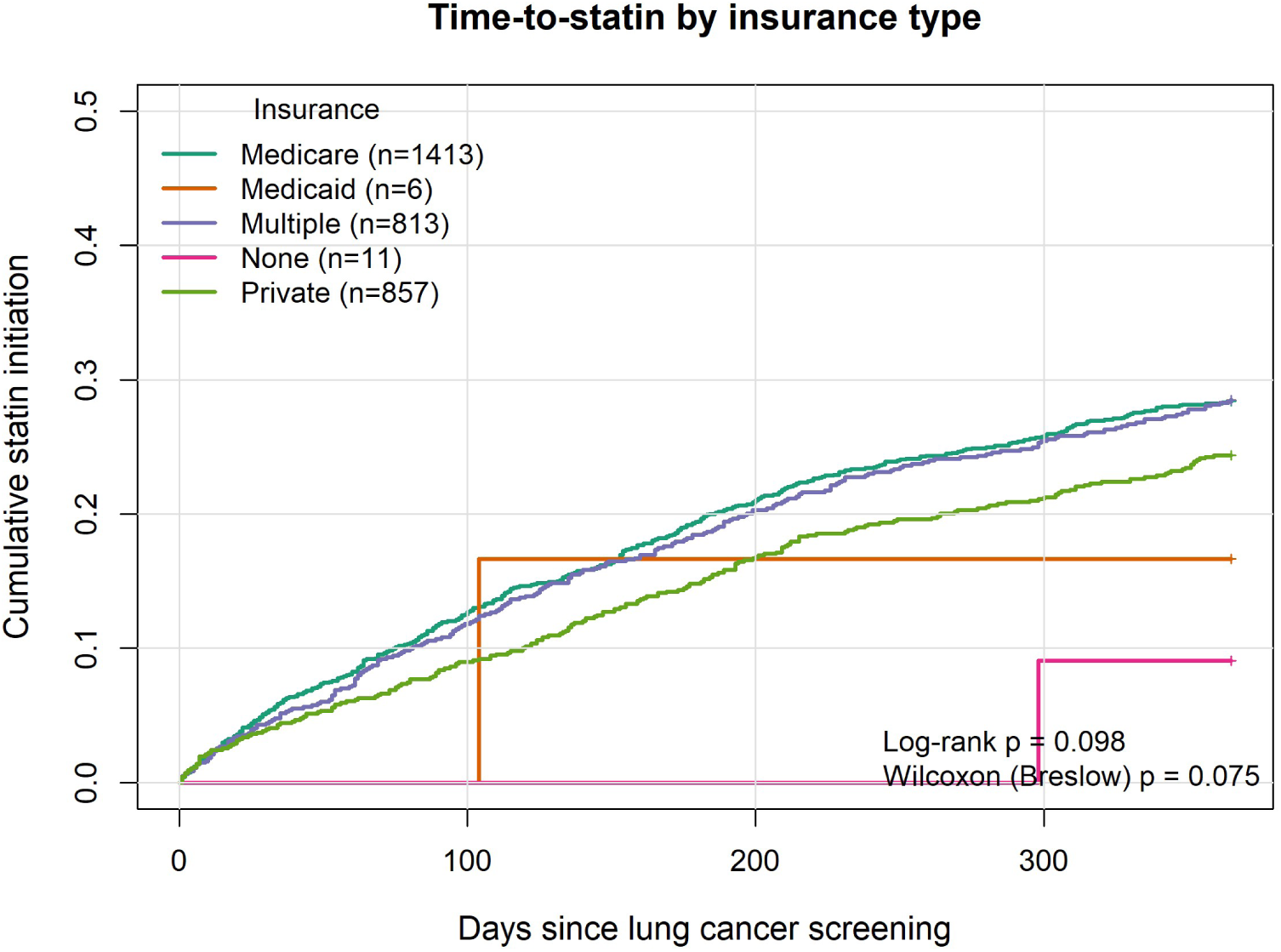
Time-to-statin prescription post-LCS by insurance type

Kaplan–Meier curves for time-to-statin initiation following LCS showed no significant differences across ADI categories (log-rank p = 0.361; Breslow p = 0.338). Survival curves for the five ADI quintiles (1 = least deprived, 5 = most deprived) overlapped, with similar trajectories throughout the follow-up period (Figure 8). No median time-to-statin initiation was reached in any group due to high censoring.

**Figure 8.**
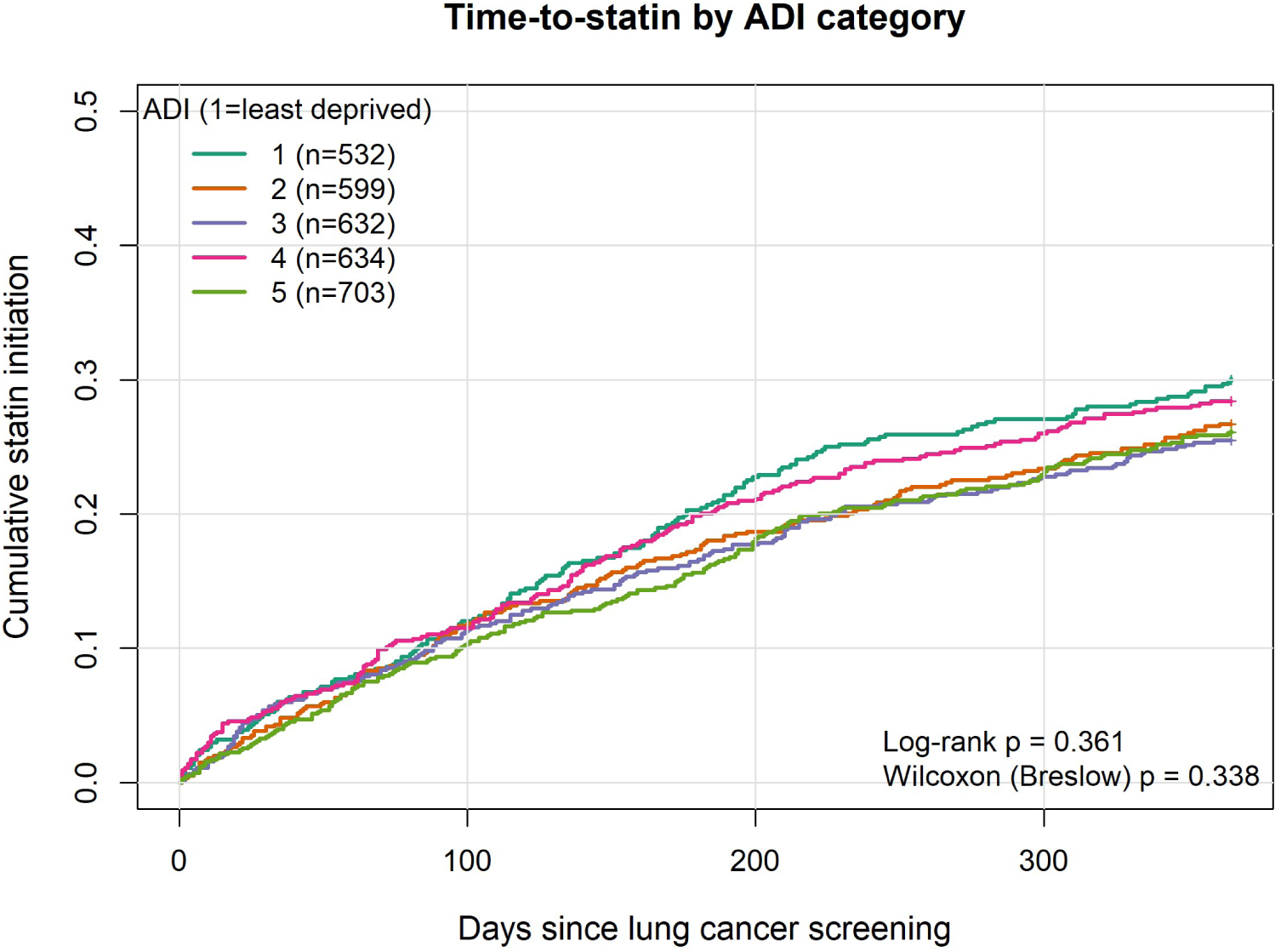
Time-to-statin prescription by ADI categories.

There was no statistically significant difference in time-to-statin initiation between males and females (log-rank p = 0.179; Breslow p = 0.248). The survival curves for males and females overlapped throughout the follow-up period, with similar rates of statin uptake over time.

There were also no statistically significant differences in time-to-statin initiation across racial groups (log-rank p = 0.398; Breslow p = 0.523). The survival curves followed a similar trajectory, and no median time-to-statin prescription was reached in any racial category due to high censoring (Figure 9).

**Figure 9.**
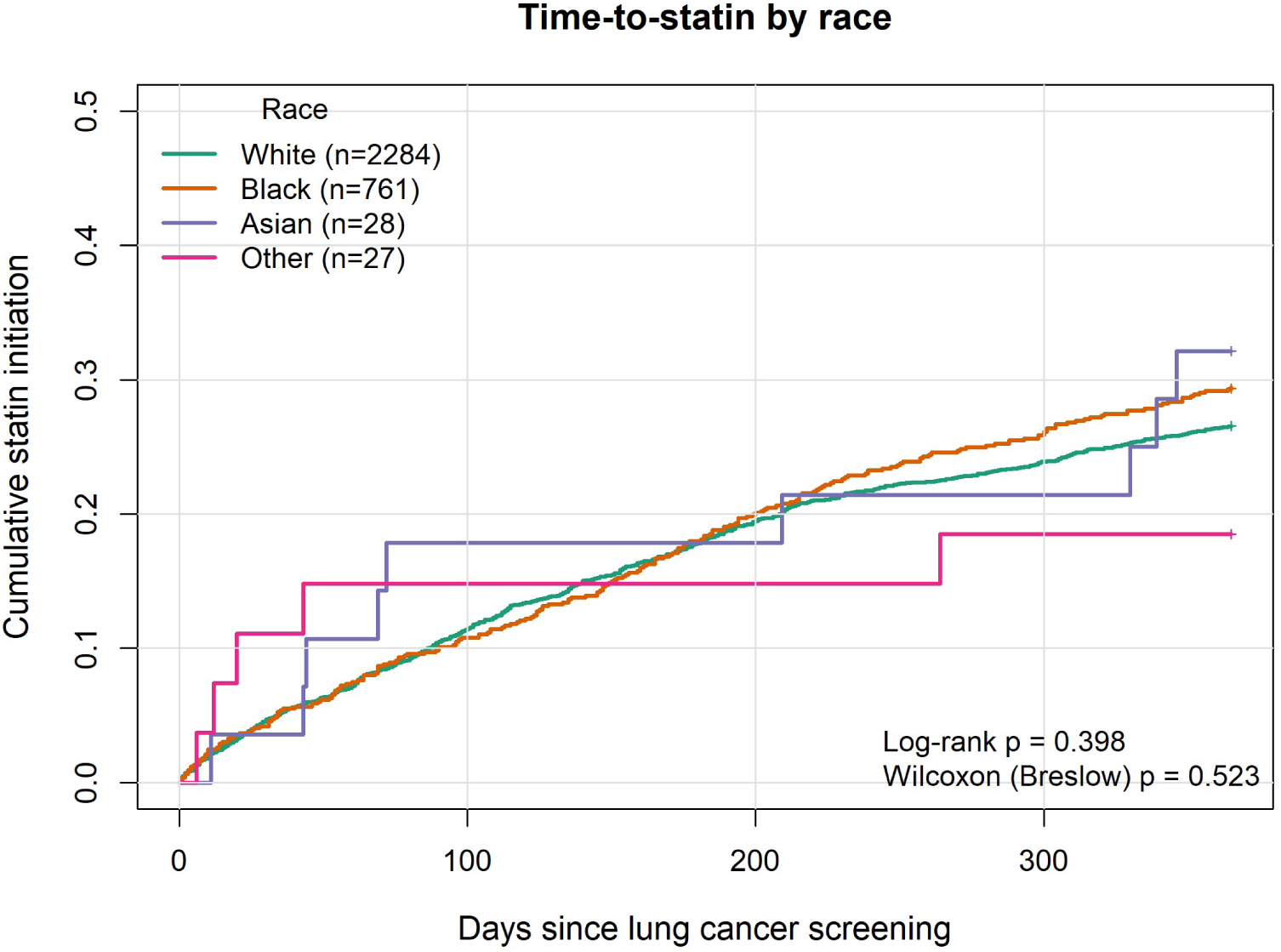
Time-to-statin prescription post-LCS

### Cox Proportional Hazards Model Results

A multivariable Cox proportional hazards model was used to evaluate predictors of time-to-statin initiation following LCS. The model included age, sex, race, BMI, smoking status, insurance type, ADI, RUCA, cardiology visit in the past year, and ASCVD risk category. Because diabetes, hypertension, and smoking are components of the Pooled Cohort Equations used to derive the ASCVD risk score, these comorbidities were accounted for through the ASCVD risk category rather than entered as separate covariates. Both unadjusted and adjusted hazard ratios are presented in Table 2.

**Table 2.**
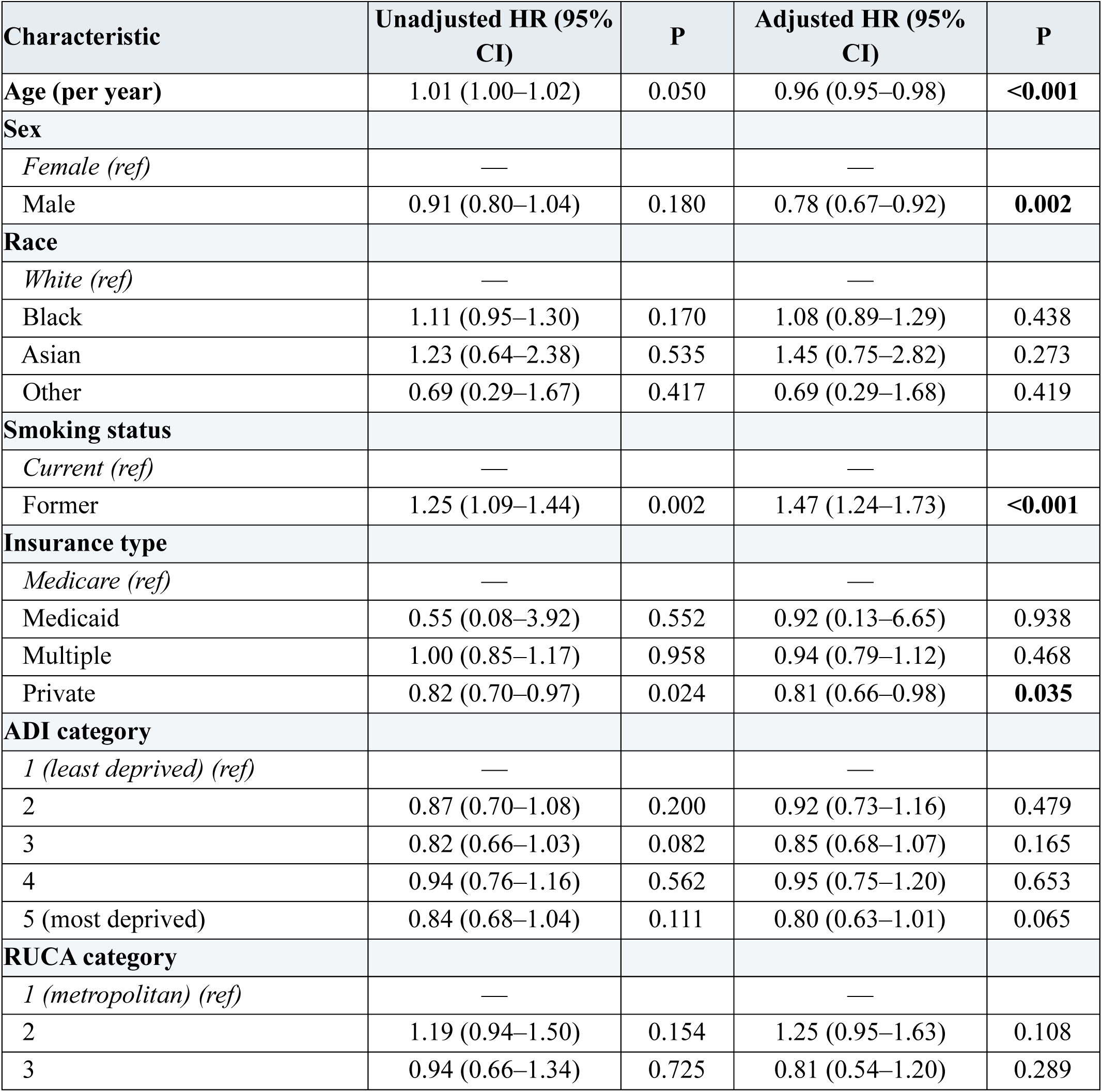
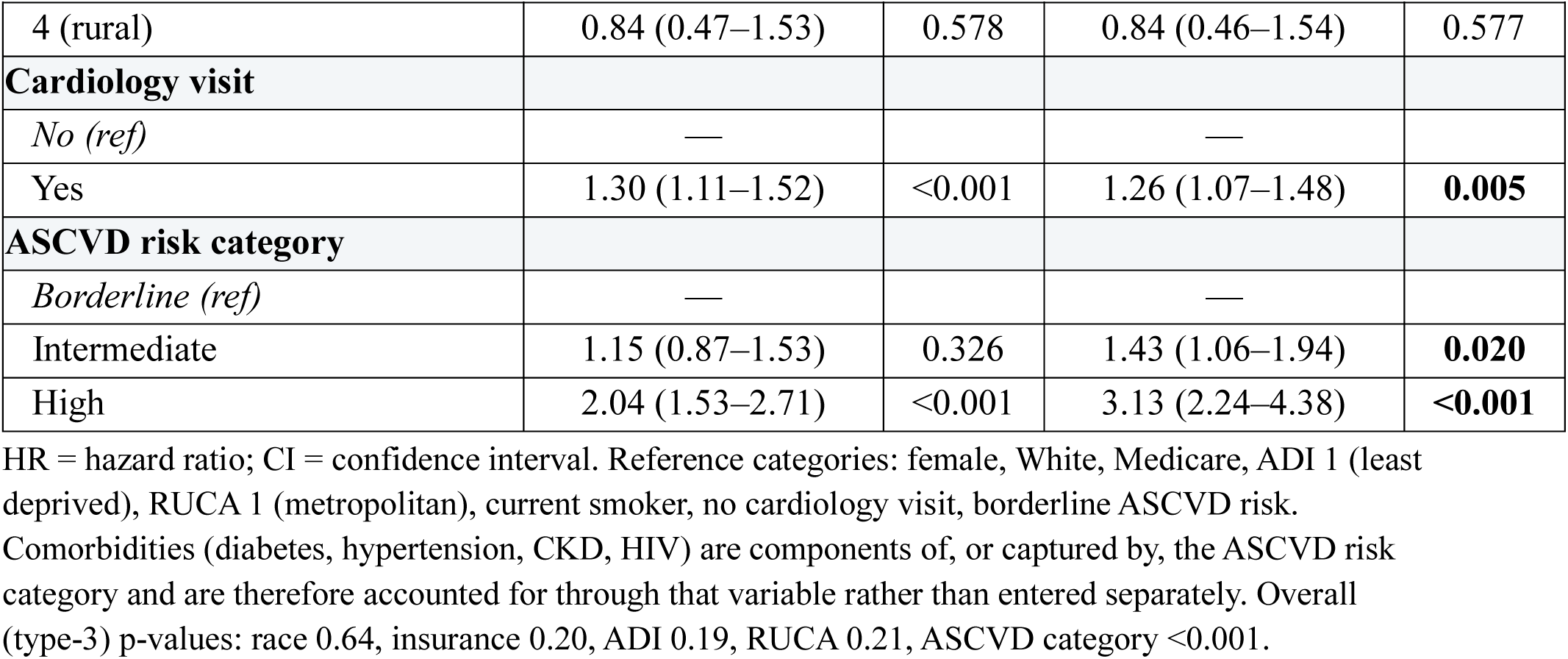
Cox Proportional Hazards Model Results.

| Characteristic | Unadjusted HR (95% CI) | P | Adjusted HR (95% CI) | P |
| --- | --- | --- | --- | --- |
| **Age (per year)** | 1.01 (1.00–1.02) | 0.050 | 0.96 (0.95–0.98) | **<0.001** |
| **Sex** |  |  |  |  |
| Female (ref) | — |  | — |  |
| Male | 0.91 (0.80–1.04) | 0.180 | 0.78 (0.67–0.92) | **0.002** |
| **Race** |  |  |  |  |
| White (ref) | — |  | — |  |
| Black | 1.11 (0.95–1.30) | 0.170 | 1.08 (0.89–1.29) | 0.438 |
| Asian | 1.23 (0.64–2.38) | 0.535 | 1.45 (0.75–2.82) | 0.273 |
| Other | 0.69 (0.29–1.67) | 0.417 | 0.69 (0.29–1.68) | 0.419 |
| **Smoking status** |  |  |  |  |
| Current (ref) | — |  | — |  |
| Former | 1.25 (1.09–1.44) | 0.002 | 1.47 (1.24–1.73) | **<0.001** |
| **Insurance type** |  |  |  |  |
| Medicare (ref) | — |  | — |  |
| Medicaid | 0.55 (0.08–3.92) | 0.552 | 0.92 (0.13–6.65) | 0.938 |
| Multiple | 1.00 (0.85–1.17) | 0.958 | 0.94 (0.79–1.12) | 0.468 |
| Private | 0.82 (0.70–0.97) | 0.024 | 0.81 (0.66–0.98) | **0.035** |
| **ADI category** |  |  |  |  |
| 1 (least deprived) (ref) | — |  | — |  |
| 2 | 0.87 (0.70–1.08) | 0.200 | 0.92 (0.73–1.16) | 0.479 |
| 3 | 0.82 (0.66–1.03) | 0.082 | 0.85 (0.68–1.07) | 0.165 |
| 4 | 0.94 (0.76–1.16) | 0.562 | 0.95 (0.75–1.20) | 0.653 |
| 5 (most deprived) | 0.84 (0.68–1.04) | 0.111 | 0.80 (0.63–1.01) | 0.065 |
| **RUCA category** |  |  |  |  |
| 1 (metropolitan) (ref) | — |  | — |  |
| 2 | 1.19 (0.94–1.50) | 0.154 | 1.25 (0.95–1.63) | 0.108 |
| 3 | 0.94 (0.66–1.34) | 0.725 | 0.81 (0.54–1.20) | 0.289 |
| 4 (rural) | 0.84 (0.47–1.53) | 0.578 | 0.84 (0.46–1.54) | 0.577 |
| **Cardiology visit** |  |  |  |  |
| *No (ref)* | — |  | — |  |
| Yes | 1.30 (1.11–1.52) | <0.001 | 1.26 (1.07–1.48) | **0.005** |
| **ASCVD risk category** |  |  |  |  |
| *Borderline (ref)* | — |  | — |  |
| Intermediate | 1.15 (0.87–1.53) | 0.326 | 1.43 (1.06–1.94) | **0.020** |
| High | 2.04 (1.53–2.71) | <0.001 | 3.13 (2.24–4.38) | **<0.001** |

HR = hazard ratio; CI = confidence interval. Reference categories: female, White, Medicare, ADI 1 (least deprived), RUCA 1 (metropolitan), current smoker, no cardiology visit, borderline ASCVD risk.
Comorbidities (diabetes, hypertension, CKD, HIV) are components of, or captured by, the ASCVD risk category and are therefore accounted for through that variable rather than entered separately. Overall (type-3) p-values: race 0.64, insurance 0.20, ADI 0.19, RUCA 0.21, ASCVD category <0.001.

Higher ASCVD risk category was the strongest independent predictor of earlier statin initiation (high vs. borderline HR 3.13, 95% CI 2.24–4.38; intermediate vs. borderline HR 1.43, 95% CI 1.06–1.94; overall p < 0.001). A cardiology visit in the prior year (HR 1.26, 95% CI 1.07–1.48, p = 0.005) and former (vs. current) smoking (HR 1.47, 95% CI 1.24–1.73, p < 0.001) were also independently associated with earlier initiation. Increasing age was associated with slower initiation (HR 0.96 per year, 95% CI 0.95–0.98, p < 0.001), as was male sex (HR 0.78, 95% CI 0.67–0.92, p = 0.002). Higher BMI was weakly associated with earlier initiation (HR 1.01 per kg/m², p = 0.043).

Race (overall p = 0.64), insurance type (p = 0.20), ADI (p = 0.19), and RUCA (p = 0.21) were not independently associated with time-to-statin initiation, although privately insured patients initiated somewhat later than Medicare beneficiaries (HR 0.81, 95% CI 0.66–0.98, p = 0.035). In a sensitivity analysis that replaced the ASCVD risk category with the individual comorbidities, diabetes (HR 1.45, 95% CI 1.24–1.69, p < 0.001) and hypertension (HR 1.43, 95% CI 1.19–1.71, p < 0.001) were themselves strong independent predictors, with CKD showing a nonsignificant trend (HR 1.17, 95% CI 0.97–1.41, p = 0.09). This confirms that the comorbidity signal and the ASCVD-category signal capture largely the same underlying risk information. The global proportional-hazards test was satisfied (p = 0.13).

## Discussion

### Summary of Key Findings

This study showed that within one year of completing LCS, only about one in four statin-eligible patients received a statin. Uptake rose with calculated cardiovascular risk, and in adjusted models a higher ASCVD risk category was the strongest independent predictor of earlier initiation, together with recent cardiology contact and former (rather than current) smoking.

Because the comorbidities that compose the ASCVD score particularly diabetes and hypertension were accounted for through the risk category to avoid double-counting, their strong crude associations are captured within it, as confirmed by a sensitivity analysis in which they re-emerged as significant predictors. Race, sex, insurance type, and area-level deprivation did not independently predict timing. The persistent gap in treatment among high-risk patients within a year of screening points to a potential structural gap in how cardiovascular prevention is delivered during the LCS encounter.

This 27.3% one-year initiation rate aligns with, or is slightly lower than, prior studies in related populations. Majeed et al. found that among LCS patients with moderate CAC, 21% remained statin-free even after the LCS finding, underscoring a missed opportunity in this setting.^7^ However, unlike incidental imaging findings that may carry more overt clinical urgency, the LCS encounter alone appears not to provoke consistent cardiovascular follow-up despite clear risk profiles. These results suggest that LCS, even when it identifies individuals at elevated ASCVD risk, is not routinely leveraged as a platform for initiating statin therapy.^7^ This represents a critical missed opportunity to address ASCVD, which remains the leading cause of death among LCS-eligible individuals surpassing lung cancer itself.^3,12^

### Lung Cancer Screening as a Missed Opportunity for ASCVD Prevention

In our cohort there was a substantial prevalence of major ASCVD risk factors, including hypertension, diabetes, and CKD. Furthermore, risk scores classified most patients as intermediate or high risk.^5^ Yet only 27.3% of statin-eligible individuals were prescribed a statin within one year of LCS. Although prescribing rose with calculated risk, the absolute uptake remained low even among high-risk patients, and the presence of risk-enhancing conditions recognized in the 2019 ACC/AHA guidelines did not translate into timely preventive action for most patients after the LCS encounter. This points to a broader issue: the lack of embedded ASCVD prevention frameworks within LCS workflows.

This represents a critical missed opportunity to address cardiovascular disease, which remains the leading cause of death among individuals eligible for LCS surpassing lung cancer itself.^12^ While the primary goal of LCS is early cancer detection, the encounter inherently targets a population at elevated ASCVD risk. Our results suggest that LCS is functioning as a siloed service, narrowly focused on cancer detection rather than integrated prevention.

This disconnect may stem from factors at multiple levels. Clinicians may prioritize cancer detection during LCS and overlook ASCVD prevention, especially without decision support or embedded risk assessments.^13^ Patients may undervalue ASCVD risk or be hesitant about statins due to perceived side effects or competing health concerns.^14^ At the facility level, fragmented workflows and lack of integration with primary care limit opportunities for follow-up.^15^ Societally, the absence of public messaging linking LCS to ASCVD prevention may further contribute to under-recognition of the broader preventive potential of these visits.

These findings mirror national patterns of underutilization of preventive therapies in high-risk populations.^6,16^ The gap is particularly pronounced in the context of LCS. Unlike breast or colorectal cancer screening, which are often bundled with broader preventive care messaging, LCS may be viewed as a single-issue intervention. Without deliberate integration of ASCVD risk assessment and follow-up into the LCS process, the encounter is unlikely to produce broader preventive benefit. This study highlights the need to rethink how LCS programs are structured and to consider them not just as cancer detection tools, but as entry points for comprehensive chronic disease prevention.^4,15,17^

### Drivers of Statin Initiation

Our study shows that statin prescribing after LCS tracked calculated cardiovascular risk. In adjusted Cox models, a higher ASCVD risk category was the strongest independent predictor of earlier initiation. In a sensitivity analysis that replaced the ASCVD category with the individual comorbidities, diabetes and hypertension were strong predictors confirming that the comorbidity signal and the calculated-risk signal reflect the same underlying information. A recent cardiology visit was also independently associated with earlier initiation, which magnifies how specialist involvement shapes care. This reflects a common pattern in preventive medicine, where clinicians act readily on both documented diagnoses and formal risk stratification once it is surfaced.^5^

Smoking status also mattered: former smokers were more likely than current smokers to have statins prescribed, despite both being guideline eligible. This may reflect provider perceptions of adherence readiness, assuming that former smokers are more motivated or engaged in self-care, or a subtle form of clinical bias, in which patients perceived as “still smoking” are deprioritized for preventive efforts.^18^ Regardless, it points to the need for consistent application of guidelines, independent of lifestyle judgment.

Although prescribing rose with calculated risk, uptake remained low in absolute terms even among high-risk patients (only 36.4% within a year). This is consistent with national data from the PALM registry^6^ and a large national cohort of patients with atherosclerosis^16^, showing that statins remain under-prescribed even among individuals meeting guideline thresholds. Several factors likely reinforce these patterns. ASCVD risk scores are not always integrated into EHR systems or surfaced during the LCS workflow, limiting their visibility at the point of care.

Providers may also face time constraints, competing priorities, or uncertainty about whether they are the “right” clinician to initiate preventive treatment.^5^ These dynamics are amplified in LCS settings, where the clinical focus is often narrow and cancer-centric.

To close this gap, ASCVD risk assessment should be integrated into LCS visits. Embedding risk calculators into LCS workflows, integrating automated prompts for statin-eligible individuals, and expanding the role of pharmacists or nurse navigators to act on these prompts could significantly improve uptake. Moreover, while CAC findings were not part of our dataset, their integration into radiology reports^4,7^ can meaningfully influence statin prescribing behavior. If LCS is to evolve into a truly preventive platform, ASCVD risk will need to be treated not as optional context, but as a co-equal clinical priority.

### Equity in Access Among Patients Engaged in LCS

While racial and socioeconomic disparities in statin use are well documented in the general U.S. population^6,9^, we did not find significant differences in statin uptake by race, insurance status, or ADI among patients in our study. Black patients were not significantly less likely to have a statin prescribed, and neither insurance type nor area deprivation was independently associated with timing. This suggests that once individuals are engaged in an LCS program, particularly within a large, integrated health system, treatment may be more standardized and less affected by demographic factors. A possible exception was a modest difference by sex, with men initiating somewhat more slowly than women.

It is possible that those who complete LCS are already more engaged in their care, which could mask disparities that occur earlier, during access or referral. While our findings are encouraging, they show only part of the care pathway. Broader barriers to screening access, follow-up, and preventive treatment likely persist and deserve further attention.

### Limitations

This study has several limitations. First, although we adjusted for a comprehensive set of clinical and sociodemographic variables, our single-institution sample may limit generalizability. Second, the high level of censoring and modest follow-up window may underestimate eventual statin uptake. Third, we were unable to capture whether CAC scores were documented or discussed during clinical visits, limiting our ability to analyze their influence on prescribing behavior directly.^4,19^ Fourth, while we used statin prescription as our outcome, we could not confirm whether patients filled or adhered to prescriptions, which may overestimate actual statin use.

### Strengths

This study has several strengths. We assessed the full spectrum of statins and non-statin alternative therapy, capturing other preventive drugs that are increasingly used in patients who are statin intolerant. We had a large dataset drawn from a large health system that provided sufficient data to examine associations with time to statin prescription. Finally, medication use was derived from documented EHR rather than self-report, avoiding recall and social desirability bias that affects survey-based estimates of statin utilization.

### Implications for Practice and Future Research

Our findings suggest that LCS programs represent a promising but currently underutilized platform for ASCVD risk reduction. Embedding ASCVD risk calculators, EHR-based prompts, or “statin check” consults into the LCS workflow could increase the efficiency and equity of preventive care. Future prospective studies should test whether co-located or team-based models of LCS and ASCVD prevention improve uptake and reduce event rates. Qualitative research exploring provider decision-making at the point of LCS interpretation may also offer valuable insights into implementation barriers.

## Conclusion

Despite targeting a population with high ASCVD risk, LCS remains an untapped opportunity for initiating statin therapy. Our findings suggest that statin prescribing tracks calculated ASCVD risk and specialist involvement, yet most high-risk, statin-eligible patients still go untreated within a year of screening. By integrating ASCVD prevention into LCS workflows, health systems have an opportunity to address two leading causes of mortality, lung cancer and heart disease in a single, streamlined encounter.

## Non-standard Abbreviations and Acronyms

ASCVD: atherosclerotic cardiovascular disease
LCS: lung cancer screening
CAC: coronary artery calcification
ACC: American College of Cardiology
AHA: American Heart Association
HER: electronic health record
ADI: Area Deprivation Index
RUCA: Rural-Urban Commuting Area
CKD: chronic kidney disease
SLE: systemic lupus erythematosus
PALM: Patient and Provider Assessment of Lipid Management
STROBE: Strengthening the Reporting of Observational Studies in Epidemiology

## Data availability

The data underlying this study contain protected health information from the Barnes-Jewish Healthcare electronic health record and cannot be shared publicly. The Institutional Review Board of Washington University approved this study under a waiver of informed consent, and the terms of that approval do not permit release of individual-level data. Aggregated data and analytic code are available from the corresponding author on reasonable request and subject to institutional approval.

## Acknowledgments

We thank the Alvin J. Siteman Cancer Center at Washington University School of Medicine and Barnes-Jewish Hospital in St. Louis, MO, for the use of the Siteman Biostatistics and Qualitative Research Shared Resource. The Siteman Cancer Center is supported in part by an NCI Cancer Center Support Grant #P30 CA091842.

